# The effect of using group-averaged or individualized brain parcellations when investigating connectome dysfunction: A case study in psychosis

**DOI:** 10.1101/2023.01.03.23284124

**Authors:** Priscila T. Levi, Sidhant Chopra, James C. Pang, Alexander Holmes, Tyler A. Sassenberg, Colin G. DeYoung, Alex Fornito

## Abstract

Functional magnetic resonance imaging (fMRI) is widely used to investigate functional coupling (FC) disturbances in a range of clinical disorders. Most analyses performed to date have used group-based parcellations for defining regions of interest (ROIs), in which a single parcellation is applied to each brain. This approach neglects individual differences in brain functional organization and may inaccurately delineate the true borders of functional regions. These inaccuracies could inflate or under-estimate group differences in case-control analyses. We investigated how individual differences in brain organization influence group comparisons of FC using psychosis as a case-study, drawing on fMRI data in 121 early psychosis patients and 57 controls. We defined FC networks using either a group-based parcellation or an individually-tailored variant of the same parcellation. Individualized parcellations yielded more functionally homogeneous ROIs than group-based parcellations. At individual connections level, case-control FC differences were widespread, but the group-based parcellation identified approximately 9% more connections as dysfunctional than the individualized parcellation. When considering differences at the level of functional networks, the results from both parcellations converged. Our results suggest that a substantial fraction of dysconnectivity previously observed in psychosis can be attributed to erroneous ROI delineation, rather than a pathophysiological process related to psychosis.

## Introduction

Psychosis is a neuropsychiatric condition that has long been thought to arise from aberrant neural connectivity, or dysconnectivity, between neuronal populations (1–4). Such dysconnectivity is often studied using a network-based approach (5), with the brains of individuals being modelled as a collection of nodes, representing discrete brain regions, connected by edges, representing inter-regional structural connectivity or functional coupling (FC). This approach has revealed extensive FC disruptions in psychosis patients, which are often characterized by a global decrease in FC upon which is superimposed more network-specific increases and decreases (1,3,4,6–11). However, the reported findings have been inconsistent, with reports of increased and decreased FC sometimes found within the same network in different samples (12–15).

Some of these inconsistencies may be explained by methodological differences in defining the nodes (brain regions of interest – ROIs) of the constructed brain networks, which is a fundamental step in network analysis that could affect the validity and interpretation of subsequent results (5,16,17). Each node should ideally represent a functionally specialized area with homogenous activity (18,19), but there is no consensus on the optimal way of parcellating the brain, meaning that investigators must rely on various heuristic methods (18,19).

The vast majority of studies in patients with psychosis have used a one-size-fits-all, group-based approach in defining distinct ROIs. A parcellation using this approach is often defined in a standardized coordinate space based on a sample average and then mapped to individual participants via a spatial normalization procedure (18). This approach fails to consider interindividual variability in functional and anatomical brain organization (20,21). Investigation of such variability with resting-state fMRI (rsfMRI) has shown that, although most cortical areas can indeed be robustly identified in every individual, their sizes and shapes vary across the population, especially when using more fine-grained parcellation methods (22). Furthermore, the topographical locations of specific areas tend to shift between individuals, sometimes across anatomical landmarks such as sulci and gyri (22), which are often used as reference points in many standard parcellations (5).

To better accommodate this individual variability, approaches have been developed to derive individualized parcellations at either the level of canonical functional networks (23,24) or cortical regions (22,25). These approaches have revealed that individual variability can considerably impact network analyses. For instance, regions assigned to one network in individual parcellations are often assigned to a different network in the group average (26), which could impact FC analysis. The use of individually-tailored parcellations yields more functionally homogeneous regions (25,27), and can improve predictions of behaviour from FC (28). Indeed, in healthy samples, individual differences in the locations of functional regions, as represented by individualized parcellation, affect predictions of fluid intelligence (28), life satisfaction (26), participant sex (29), and performance in reading and working memory tasks (25). Moreover, some estimates indicate that up to 62% of variance in network edge strength (i.e., FC values) can be explained by the spatial variability of defined regions (26). These findings suggest that clinically important relationships may be masked when using a group-based parcellation.

A particularly salient point in clinical studies, such as those of schizophrenia, is that standard brain atlases have been derived from healthy participants, which may not adequately capture the characteristic properties in the brain organization of patients (30,31). Patient-specific individual variability in functional organization can influence the results of brain network analyses. Indeed, one study has found that slight displacements of a seed region in the thalamus can lead to significant differences in disorder-related dysconnectivity (32), emphasizing the importance of a valid and consistent node definition.

One strategy to develop individualized parcellations is to adjust the borders of a group-based template for each individual participant according to pre-defined functional criteria. For instance, Chong et al. (27) developed a Bayesian algorithm (called Group Prior Individualized Parcellation – GPIP) that uses a group-based template as a prior to find an optimal corresponding parcellation on individual brains using individual’s FC data. The group-based prior ensures that the same regions are mapped in each individual, while updates to the individualized prior account for variability in the shape and size of each parcellated region. Chong et. al. (27) has shown that this method yields parcellated regions with increased intra-regional functional homogeneity and reduced variance in connectivity strength between individuals (27). Here, we used this approach to compare FC disruptions observed in people with early psychosis using analyses that rely on either a group-based or individualized parcellation. The parcellation algorithm (27) allowed us to match all brain regions across participants while accounting for individual variability. Our analyses were conducted using the high-quality, open-access data provided by the Human Connectome Project - Early Psychosis (33,34) (HCP-EP) resource. We tested two competing hypotheses of how individual variability contribute to apparent FC disruptions in psychosis. Under one hypothesis, a failure to consider individual variability may lead to erroneous regional parcellations, adding noise to the analyses and reducing statistical power for detecting valid group differences. In this case, we expect to see fewer differences between patients and controls when using the group-based parcellation compared to individualized parcellation. Alternatively, FC differences between groups may be largely driven by variations in the underlying organization of each individual’s brain, rather than reflecting specific differences in FC. In this case, we expect to see more differences using the group-based parcellation.

## Results

Here, we present results obtained using group-level cortical parcellations provided by Schaefer et al. (30) as the basis for our analysis, focusing on the 100-region parcellation (s100). To ensure that our results are robust to the number of regions, we repeated our analysis using the 200-region variant (s200) and after applying Global Signal Regression (GSR). Results obtained using the s200 atlas, and results for both atlases after GSR, can be found in the supplementary material and are largely consistent with the primary results reported in the following sections.

### Spatial and functional properties of group-based vs individualized parcellation

Figure 1 shows examples of individualized parcellations generated for three individuals compared with the original group-based s100 atlas. The individualized parcellation algorithm preserved the same regions for every individual but shifted their borders and changed their shapes and sizes to accommodate for individualized variations in brain organization. Indeed, on average, 42.56% (*SD* = 2.37) of vertices were reallocated to a different region as a result of the individualized parcellation algorithm, highlighting the considerable variability of cortical functional organization between individuals. Figure 2a shows the proportion of edges that were relabelled in controls *M*(*SD*) = 43.28% (2.34) and in patients *M*(*SD*) = 42.20% (2.31). The difference between the two groups was small but statistically significant (*t*(165) = 2.824, *p* = 0.005, 95%*CI* = [0.003, 0.018]).

**Figure 1.**
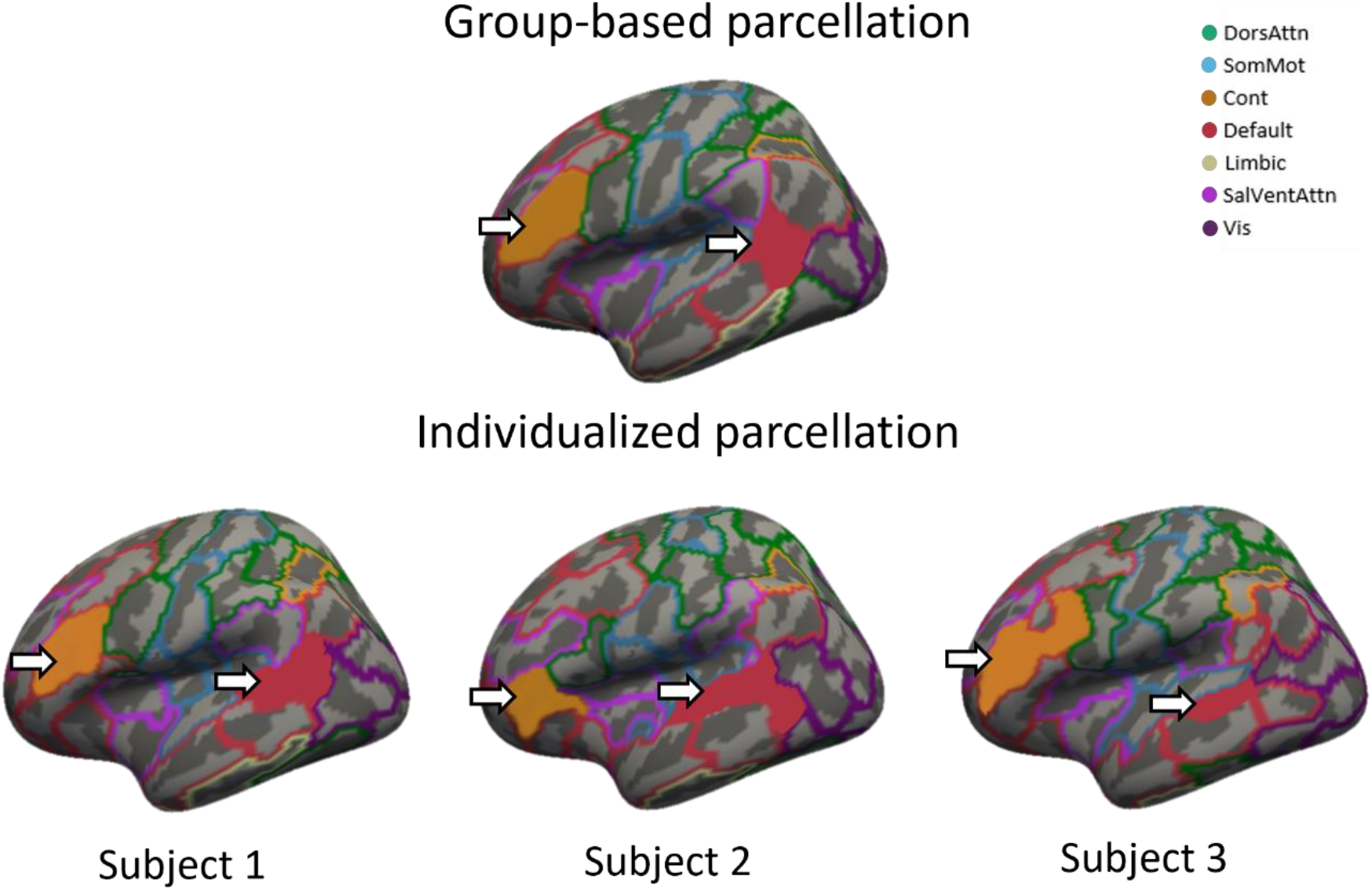
Differences in parcel boundaries between group-based and individualized parcellation. The images show different parcellations overlayed on the inflated fsaverage5 template surface of the left hemisphere, with 20,484 vertices. The top image shows the group-based parcellation, which was used as a starting point for the individualized parcellation algorithm. Colors correspond to the seven canonical functional networks that are used to group parcels in the atlas (23). The bottom three images show individualized parcellations for three different subjects after 20 iterations of the GPIP algorithm. The region shaded in orange corresponds to region 1 in the lateral prefrontal cortex of the control network for all parcellations. The region shaded in red corresponds to region 1 in the parietal lobe of the Default Mode Network. The same regions are present in all individuals, but their locations, sizes and shapes show considerable variability. DorsAttn – dorsal attention network; SomMot – somatomotor network; Cont – control network; Default – default mode network; Limbic – limbic network; SalVentAttn – salience/ventral attention network; Vis – visual network.

**Figure 2.**
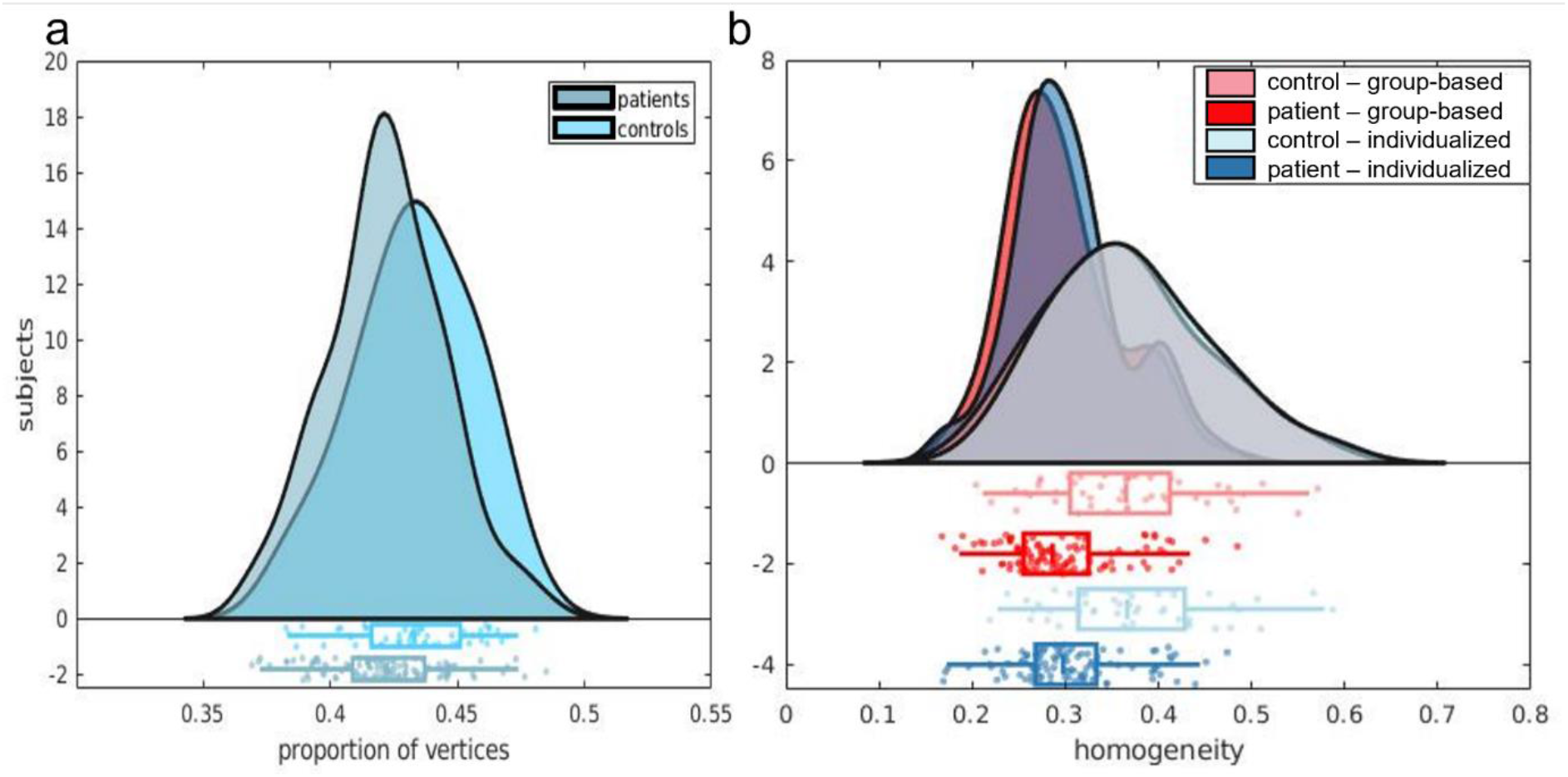
Spatial and functional properties of group-based vs individualized parcellation. Panel **a** shows the proportion of vertices reallocated by the individualized parcellations for controls (*M*(*SD*) = 0.433(0.023)) and for patients (*M*(*SD*) = 0.422(0.023)). Panel **b** shows the distribution of homogeneity scores per subject. Controls produced more homogenous parcels in both individualized (*M*(*SD*) = 0.372(0.08)) and group-based parcellations (*M*(*SD*) = 0.364(0.09)) than patients (*individualized M*(*SD*) = 0.304(0.06)), (*group* − *based M*(*SD*) = 0.297(0.06)).

We next compared the average functional homogeneity of the group-based and individualized parcellations. Functional homogeneity was measured out of sample, on functional scans from run 2 with parcellations generated for scans from run 1. In controls, the mean homogeneity was 0.364 (*SD* = 0.09), and 0.372 (*SD* = 0.08) for the group-based and individualized parcellations, respectively. In patients, the mean homogeneity was 0.297 (*SD* = 0.06) and 0.304 (*SD* = 0.06) for the group-based and individualized parcellations, respectively (figure 2b). A two-way mixed ANOVA revealed that mean homogeneity was higher for the individualized parcellation (*F*(149) = 54.81, *p* < 0.0001) and higher in controls compared to patients (*F*(149) = 30.91, *p* < 0.0001), with no interaction between parcellation type and diagnostic group (*F*(149) = 0, *p* = 0.898).

### Unthresholded edge-level group differences in FC

Following exclusion of regions with poor signal (see Methods) the final networks examined comprised 85 regions. The FC matrices resulting from both parcellation methods were positively correlated, with correlations ranging between 0.679 and 0.898 (median = 0.794) across participants (Supplementary Material figure 4).

Figure 3a shows the distribution of *t*-statistics across edges, comparing FC between patients and controls estimated using either the group-based or individualized parcellation. Both distributions have predominantly positive values, consistent with evidence of widespread hypoconnectivity in patients compared to controls. The distribution for the group-based approach is shifted further to the right, indicating that larger group differences are detected with this method, on average. The difference in the means of the distributions was statistically significant, as calculated with a Wilcoxon signed-rank test (*p* < 0.0001). Given the higher functional homogeneity of the individualized parcellation, this result suggests that the group-based parcellation overstates FC differences between patients and controls.

**Figure 3.**
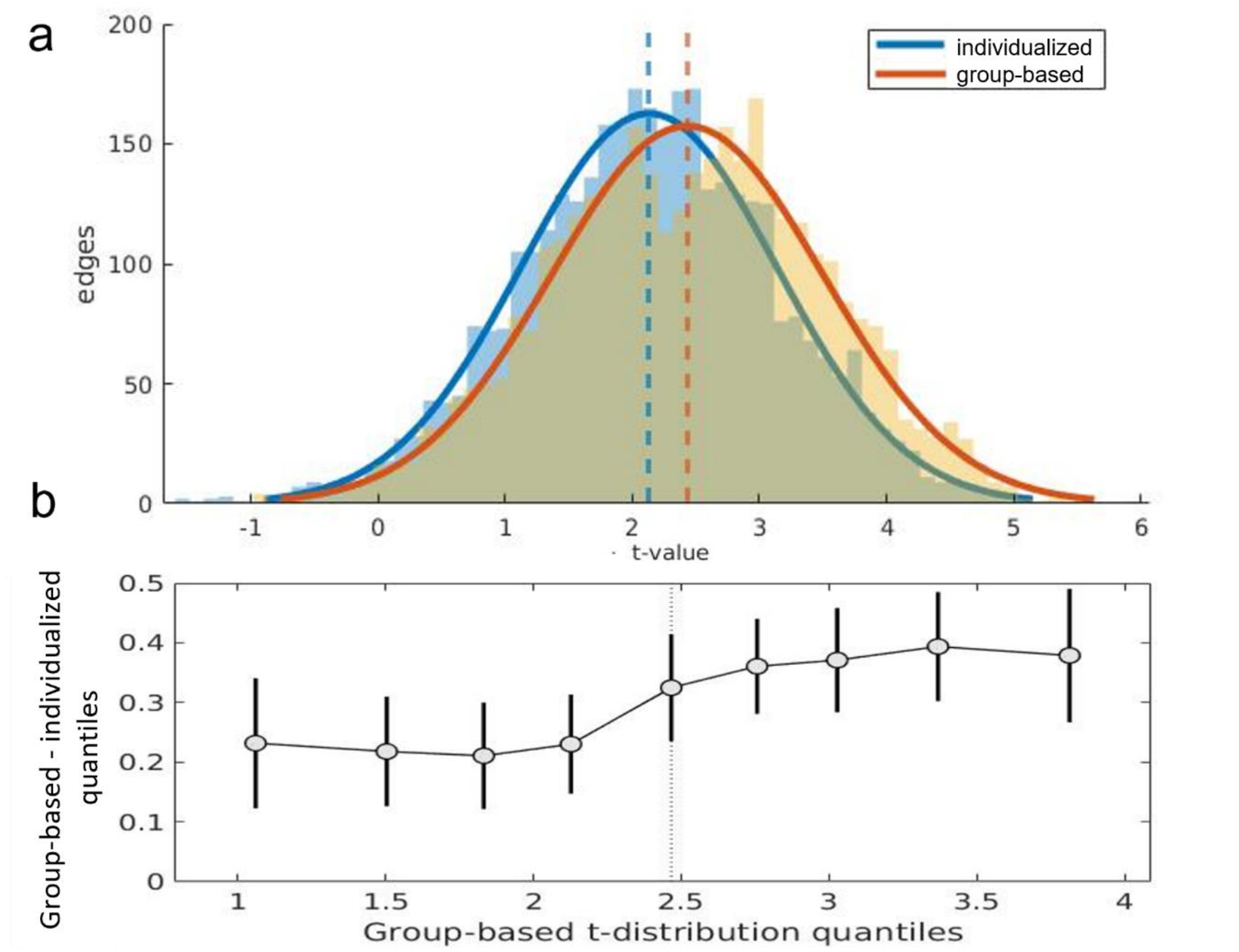
Edge-specific case-control differences in FC depend on parcellation type. **a** Distributions of *t*-values quantifying FC differences between patients and controls at each edge and for each parcellation type. Positive t-value indicate a greater FC value in controls than in patients. **b** Shift function (35) for the two t-distributions. Each circle represents the difference between the borders of each decile of both distributions as a function of the deciles in the group-based distribution. The bars represent the 95% boot-strap confidence interval associated with the difference.

The *t*-matrices obtained using the group-based and individualized parcellations were only moderately correlated (*r* = 0.56), suggesting that the two approaches can lead to substantially different conclusions about the edges that are the most affected in patients with psychosis. These effects of parcellation type were consistent across the full extent of the *t*-distributions, as indicated by the shift function, which compares differences between distributions at each decile. This analysis showed a significantly higher value in every decile of the group-based parcellation, when compared to the individualized parcellation, with the 95% CI never crossing zero (figure 3b). There was, however, a more pronounced effect of parcellation type on edges associated with larger case-control differences in FC relative to those with smaller case-control differences, as can be seen by the greater shift observed in the right tail of the distribution relative to the left (figure 3b). This result implies that variations in parcellation type are more likely to influence the edges that are significantly different between patients and controls.

### Thresholded edge-level group differences in FC

We used the Network Based Statistic (NBS) for inference on the edge-specific *t*-statistics (36). The NBS identified a single connected component with significant FC differences between patients and controls using both the group-based (*p* = 0.0002) and individualized parcellations (*p* < 0.0001). Out of 3,570 possible connections, the group-based and individualized parcellations resulted in components comprising 2,712 edges and 2,461 edges respectively (figure 4a-b). Thus, the group-based approach implicated approximately 9% more dysconnected edges. The binary edge matrices defining these components were positively correlated (*r* = 0.795) and both components had a total of 643 edges that differed from each other. There was also some variation in the regional affiliation of the edges. For example, figures 4c-d show that the insula has a high dysconnectivity degree in both group-based and individualized parcellations, but that the former approach implicates more insula sub-regions. Furthermore, the right medial prefrontal cortex shows a low degree in the individualized parcellation but not in the group-based parcellation.

**Figure 4.**
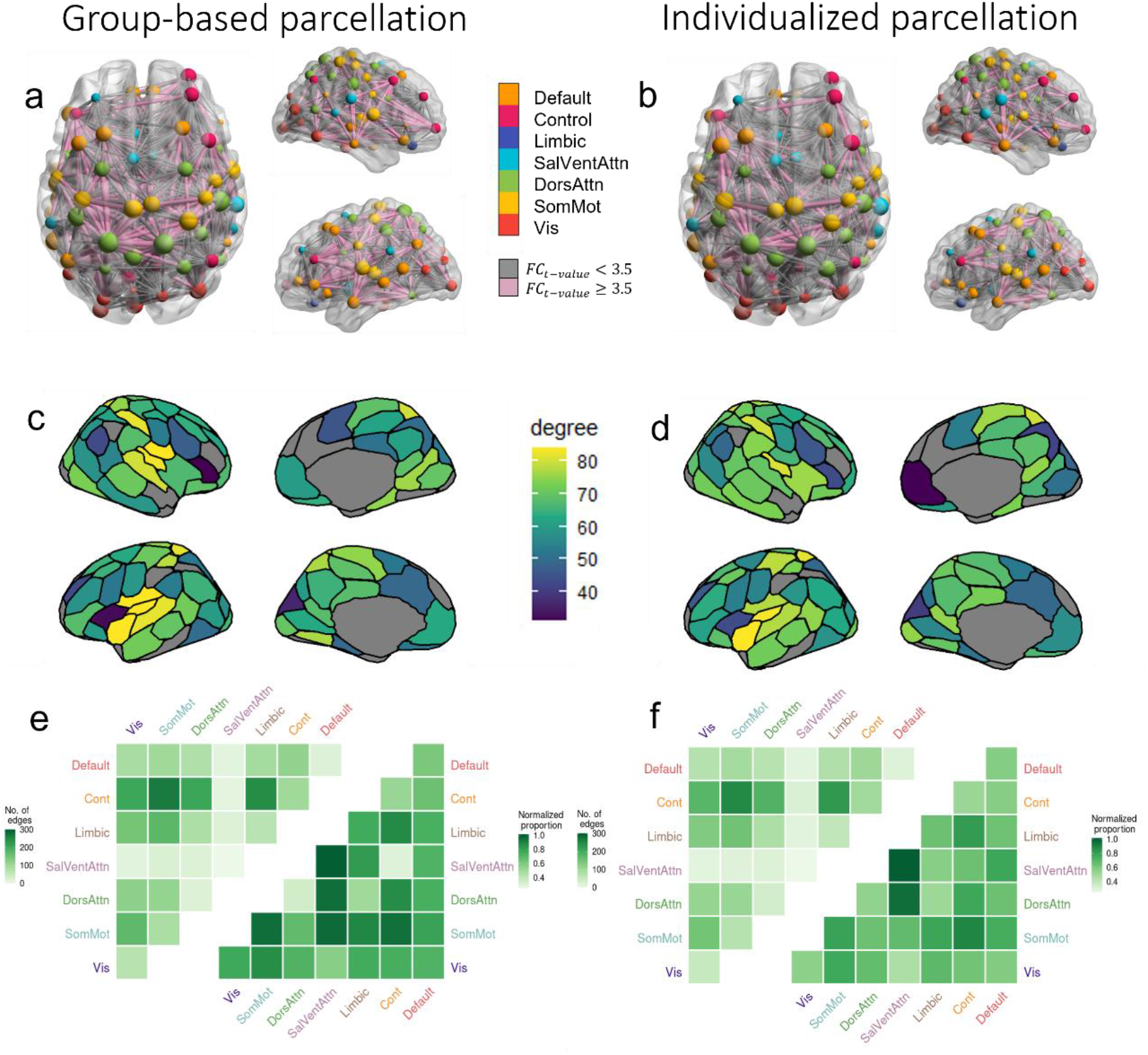
Edge-level regional and network-level case-control FC differences according to parcellation type. Panels **a** and **b** show the specific edges comprising the NBS components obtained with the group-based and individualized parcellations, respectively, with nodes colored according to network affiliation and sized by degree. Edges are sized by strength of dysconnectivity. Edges associated with a t-value < 3.5 are represented by grey lines and those associated with a t-value ≥ 3.5 are represented in pink. The images were created using the software BrainNet Viewer (38). Panels **a, c**, and **e** results from the group-based parcellation. Panels **c** and **d** show the degree of each region in the NBS component for the group and individualized parcellations, respectively. The left most triangle of each matrix in panels **e** and **f** shows the total number of NBS component edges (raw counts) falling within and between seven canonical networks. The right most triangles show the same data normalized for network size, i.e. the total number of possible connection within or between networks (normalized counts). DorsAttn – dorsal attention network; SomMot – somatomotor network; Cont – control network; Default – default mode network; Limbic – limbic network; SalVentAttn – salience/ventral attention network; Vis – visual network.

### Effects of variations in parcel size

A challenge of using individualized parcellations is that the ROIs can vary in size across individuals, which may bias estimates of FC differences between groups. We therefore examined changes in parcel size resulting from the individualization algorithm, as quantified by the number of vertices in each parcel. On average, parcels changed by 56.9 (SD = 42.0) vertices across patients and 57.5 (SD = 41.9) across controls, with no significant difference between the two groups (*p* = 0.889) (Supplementary Materials figure 8a). There was also no significant difference in size difference between patients and controls for any of the parcels (i.e., all *p* > .05, Bonferroni-corrected). We next correlated the differences in parcel size between patients and controls with differences in node degree within the NBS network and mean edge dysconnectivity, given by the mean *t*-value of edges attached to each node for the case-control comparison (Supplementary Materials figure 8b-c). Both correlations were moderately positive (*r* = 0.217 and *r* = 0.277, respectively), suggesting that large group differences in FC were identified in regions that show greater size differences between patients and controls.

### Network-level group differences in FC

Having demonstrated that the choice of a parcellation strategy can influence both edge- and region-level inferences about FC disruptions in psychosis, we next examined whether parcellation type affects the specific networks that are considered to be dysfunctional. We therefore examined the proportion of edges within the NBS network that fell within and between each of 7 canonical functional networks (37). Considering the raw number of affected edges across both parcellation approaches, the control network was the most impacted in patients with psychosis, with over 1,000 dysconnected edges, particularly those linking the control and somatomotor networks (figure 4e-f).

By comparison, normalized counts, which is adjusted for the total number of possible edges within or between pairs of networks, suggested a more equal and widespread distribution of FC disruptions across networks. Both the raw count (*r* = 0.995) and normalized matrices (*r* = 0.723) were strongly correlated across the two parcellation methods. These findings indicate that while parcellation method can influence the specific edges that are identified as dysconnected, these edges generally fall within or between the same canonical networks.

## Discussion

Several studies have reported functional brain dysconnectivity in psychosis. A fundamental step in such analyses involves defining a priori ROIs to serve as nodes in the network analysis, which are typically derived from standard parcellation atlases generated from a population or group average template. Here, we asked whether the failure of such an approach to account for individual differences in brain functional organization can bias estimates of case-control differences in FC. Such a bias could manifest as either an under-estimation of the true extent of network dysfunction (due to noisy FC estimation caused by inaccurate ROI delineations) or as an inflated estimate of the true dysfunction (due to FC differences being attributable to ROI misalignment). Our findings indicate that group-based parcellations inflate estimates of FC differences in psychosis. Moreover, the use of individualized parcellations, while yielding a generally consistent pattern of findings, leads to substantially different conclusions about the specific edges and regions most affected by the disorder, although inferences at the network level were robust to parcellation variations. Together, our findings suggest that the use of individualized parcellations can yield more refined maps of brain dysconnectivity in psychosis and, by extension, other disorders.

### Individualized parcellations yield more functionally homogeneous regions

The individualized parcellations resulted in nearly half (over 40%) of vertices being assigned to regions that differed from the group-based atlas, as per prior work (27). This finding reiterates how group-based parcellations can result in a substantial misspecification of regional borders in individuals and highlights the high degree of variance present in the topographical organization of functional areas. The higher functional homogeneity of the individualized parcellations supports its improved validity, although the increment was small (2.4%), which is consistent with past reports (25,39). Regional homogeneity was also marginally (2.3%) higher in controls compared to patients. This differential improvement in homogeneity was expected, as the starting point for the GPIP algorithm was the Schaefer atlas (30), which was derived from a sample of people with no psychiatric disorders. Defining an initial group atlas in patients would better account for differences in cortical functional organization caused by schizophrenia, but it would complicate comparisons between groups because of the requirement to have consistently defined nodes in both patients and controls. Since most case-control studies use data obtained in healthy individuals to establish a normative benchmark for measures acquired in patients (3,10,40), we relied on the Schaefer parcellation in our analysis. Future work could develop methods to better capture variations in functional organization associated with schizophrenia.

### Individualized parcellations lead to more conservative estimates of case-control FC differences

Although widespread decreases in FC in patients with psychosis were identified using both parcellation approaches, the magnitude of the differences was greater in the group-based parcellation compared to individualized parcellation. Notably, the shift function analysis indicated that differences between the two parcellation approaches were greater for edges associated with large case-control differences. These edges are precisely the ones that are most likely to be declared as statistically significant following the application of some thresholding procedure. Accordingly, comparison of NBS results revealed a 9% reduction in the size of the dysfunctional component identified using the group-based parcellation. Given the higher functional homogeneity, and thus validity, of the individualized parcellation, these results support the hypothesis that at least part of the group differences identified in past studies in schizophrenia samples do not reflect actual differences in inter-regional FC but instead result from inaccurate ROI boundaries caused by a failure to account for individual differences in functional organization. These findings imply that individualized parcellations can yield more refined and precise estimates of FC differences in case-control studies.

### Parcellation type affects the location of FC differences in edges and regions, but not networks

While widespread decreases in FC were apparent in patients with psychosis using both parcellation methods, the specific edges affected varied considerably. The NBS components of both group-based and individualized parcellations showed differences in 643 edges (i.e., 23% of the total identified with the group-based parcellation). Examining the regions most affected by quantifying the node degrees of the NBS components resulted in broadly similar patterns, but there were some notable differences in medial frontal and insula regions. These findings suggest that conclusions about the specific edges and regions affected by psychosis can vary considerably depending on the parcellation method used. In contrast, inferences at the network level were largely consistent across the two parcellation approaches, indicating that coarse-grained localizations of FC differences are robust to this methodological choice. This could be attributed to network-level inference effectively reducing the dimensionality, minimizing the nuances of more fine-grained individual variations.

### Limitations

To minimize the computational cost, we used fsaverage5, a surface mesh with a relatively low number of vertices. Since GPIP parameters depend on the number of vertices of the mesh, future work could investigate the impact of different surface mesh resolutions and whether the differences observed here apply at different mesh resolution.

A proportion of patients in our sample were medicated, and recent evidence has shown that anti-psychotic medication can impact FC, even after only 3 months of use (10). However, given that most samples examined in past research are also medicated, our sample is directly comparable to the broader literature. We also emphasize that this study is not focused on identifying the specific nature of FC disturbances associated with psychosis but instead concentrates on how parcellation type affects FC differences in the same patients. In this context, medication exposure was constant across our main contrast of interest (parcellation type), meaning that it cannot explain the differences that we focus on here. The same reasoning applies to the clinical heterogeneity of the patient sample, which comprised people diagnosed with both affective and non-affective psychoses. Future work could use individualized parcellations to delineate FC differences more precisely between distinct patient subgroups.

### Conclusion

Our findings indicate that traditional reliance on group-based parcellations may inflate case-control differences in FC. The use of individualized parcellations yields a more conservative and refined understanding of brain network disruptions in psychotic and other disorders.

## Study participants

### Methods

All data for this study were collected as part of the Human Connectome Project – Early Psychosis (HCP-EP) study, which is an open-access collection aiming to generate high-quality imaging data in early psychosis patients and healthy controls (33). This study includes high-resolution structural and functional Magnetic Resonance Image (MRI) data from 121 patients with early psychosis (74 males) and 57 healthy individuals (37 males). Demographic information is provided in Table 1. Data collection by HCP-EP has been approved by the Partners Healthcare Human Research Committee/IRB, and comply with the regulations set forth by the Declaration of Helsinki (41).

**Table 1.**
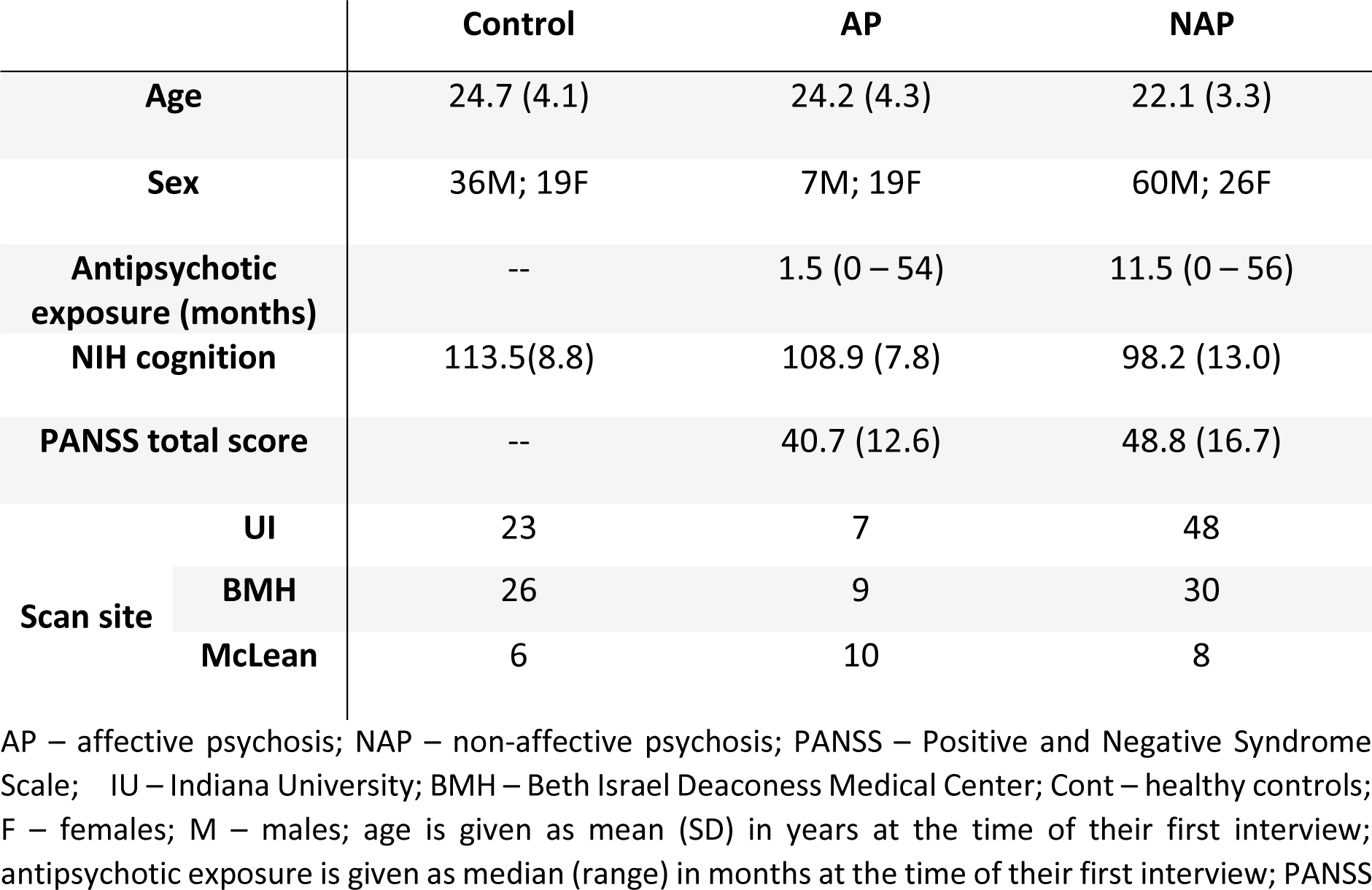
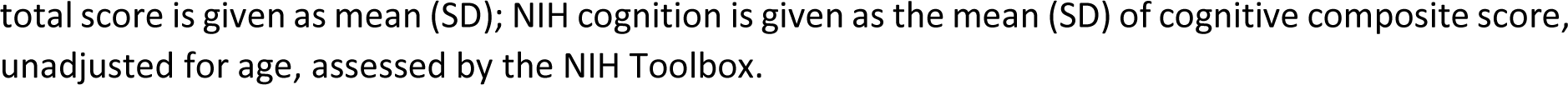
Demographic details.

The patient group was comprised of outpatients with psychosis, meeting criteria for affective or non-affective psychosis, according to the DSM-5, who were within the first five years of onset of symptoms. Patients were recruited by four clinical programs: Beth Israel Deaconess Medical Center (BMH) – Massachusetts Mental Health Center (BIDMC-MMHC), Prevention of and Recovery from Early Psychosis (PREP) Program; Indiana University Psychotic Disorders Program, Prevention and Recovery for Early Psychosis (PARC); the McLean Hospital, McLean On Track; and Massachusetts General Hospital, First Episode and Early Psychosis Program (FEPP) (33). Imaging took place in three of these sites.

The control group included volunteers that did not present with anxiety disorders and/or psychotic disorders, had no first-degree relative with schizophrenia spectrum disorder, were not taking psychiatric medication at the time of the study, and had never been hospitalized for psychiatric reasons. All participants were aged between 16 and 35 years old (mean = 23, SD = ±3.9) at the time of the study (Table 1). A total of 11 subjects were excluded due to poor data quality, as detailed below, leaving a final sample of 55 (36 male) controls and 112 (67 male) patients.

### Data Acquisition

The participants were recruited from four locations were scanned at three sites: BMH; Indiana University; and McLean Hospital, using Siemens MAGNETOM Prisma 3T scanners. The acquisition parameters between the three sites were harmonized and followed the widely used HCP protocol (33,42). The project collected whole brain T1-weighted MRI (T1w), T2-weighted MRI (T2w), diffusion MRI, spin echo field maps with Anterior to Posterior (AP) and Posterior to Anterior (PA) phase encoding (PE) directions - and four resting-state functional MRI (rsfMRI) sessions. The current study uses the T1w and T2w images, the spin echo field maps, and the first two runs of the rsfMRI scans. A 32-channel head coil was used at BMH and Indiana University. A 64-channel head and neck coil, with neck channels turned off was used at McLean Hospital. Real-time image reconstruction and processing was performed for quality control and scans with detectable problems were repeated (33).

### Structural MRI acquisition parameters

Acquisition parameters followed HCP standards. T1w images were obtained using a magnetization-prepared rapid gradient-echo (MPRAGE), with 0.8 mm isotropic spatial resolution echo time (TE) = 2.22 ms, repetition time (TR) = 2400 ms, and field of view (FoV) = 256 mm. T2w images were acquired following a 3D-SPACE sequence, with 0.8 mm isotropic spatial resolution, TE = 563 ms, TR = 33200 ms, and FoV = 256 mm (43).

### Functional MRI acquisition parameters

The present study mainly utilized the first rsfMRI run (with anterior to posterior phase encoding). The second run (with posterior to anterior phase encoding) was used to validate the parcellation with out-of-sample analysis of within parcel homogeneity. Scans were acquired for a length of 6.5 minutes, resulting in a total of 420 volumes; the first 10 volumes were removed prior to the dataset release. Images have an isotropic spatial resolution of 2 mm, TE = 37 ms, TR = 800 ms, and FoV = 208 mm. A multi-band acceleration factor of 8 was used to improve spatial and temporal resolution (43).

### Structural and Functional Image Analysis

#### Raw Image Quality Control

All raw structural and functional images were first visually inspected for large artefacts and distortions. Images were then put through an automated quality control pipeline (MRIQC) (44) which computes 15 image quality metrics for each scan with the purposes of identifying outliers warranting closer inspection. At this stage, three subjects were excluded for missing or unusable structural images.

Head motion is a major source of noise in fMRI signals. Its effects remain present even after volume realignment and can introduce systematic bias in case-control studies when not strictly controlled (45,46). Head motion during the fMRI scan was estimated using frame-wise displacement (FD), which is a summary measure of the movement of the head from one volume to the next (46). For each scan, FD was calculated according to the method described by Jenkinson et al. (47) and the resulting trace was band-pass filtered and downsampled to account for the high sampling rate of the multiband fMRI acquisition (48). Subjects were excluded if they met at least one of the following stringent exclusion criteria: scans had a mean filtered FD greater than 0.25 mm; more than 20% of frames were displaced by more than 0.2 mm; or any frame was displaced by more than 5 mm. These criteria have previously been shown to effectively mitigate motion-related contamination in fMRI connectivity analyses (46). In total, 11 subjects (2 controls) were excluded for excessive head movement in the scanner.

#### Image Preprocessing

T1w images were processed using FreeSurfer version 6.0.1 (49) to generate cortical surface models for each participant. Surfaces were visually examined for inaccuracies and distortions. The fMRI data were processed according to the Minimal Preprocessing Pipeline for HCP data (34). The pipeline adapts steps from FMRIB Software Library (FSL) and FreeSurfer to account for greater spatial and temporal resolution and HCP-like distortions resulting from acquisition choices such as multiband acceleration (34). Briefly, images were skull stripped by the brain extraction tool (BET) (50) of FSL, which removes non-brain matter from the image. Skull stripped T1w, T2w, and fMRI were aligned using FMRIB’s Linear Image Registration Tool (FLIRT) (47,51). Spin Echo EPI field maps with opposite phase encoding directions were used to estimate spatial distortion caused by magnetic field inhomogeneities, with corrections applied using FSL’s “topup” (52) and FLIRT. This process was fine-tuned and optimized using FreeSurfer’s BBRegister (53). Furthermore, bias field correction was performed on structural images to remove gradients of voxel intensity differences, following the HCP pipeline (34). The fMRI volumes were realigned to the first volume for each participant using FLIRT. The fMRI data were then co-registered to their structural image, and the structural image was non-linearly normalized into standard Montreal Neurological Institute (MNI) ICBM152 space (54) using FLIRT and FMRIB’s nonlinear image registration tool (FNIRT) (55). The resulting transform was then applied to the functional data.

#### fMRI Denoising

The functional data were denoised using Independent Component Analysis (ICA)-based X-noiseifier (FIX), which decomposes the data into spatially independent components and uses machine learning to label each resulting component as either signal or noise (56,57). The preprocessed fMRI timeseries were then regressed against the estimated noise component signals and the residuals were retained for further analysis. Component decomposition was performed using Multivariate Exploratory Linear Optimized Decomposition into Independent Components (MELODIC) (56,57). HCP’s training set – HCP_hp2000, which includes pre-trained weights to classify independent components, was used as the training set for the algorithm. A temporal high-pass filter of 2000 was applied to remove low-frequency signal drifts (34). Following HCP’s guidelines (34), a lenient threshold component labelling in FIX was used (th=10), regressing out the noise components while controlling for the signal components. The accuracy of the labels was manually verified. The analyses were repeated after applying Global Signal Regression (GSR), which removes widespread signal fluctuations associated with respiratory variations (58,59) (see Supplemental Material).

#### Surface Registration

The processed images in MNI volume space were resampled to each individual’s cortical surface, as generated by FreeSurfer, and then registered to the fsaverage5 template using a surface-based registration algorithm (49,60). fsaverage5 is a standard template generated by FreeSurfer, the resulting surface mesh comprises a total of 20,484 vertices.

#### Parcellations

We used group parcellations provided by Schaefer et al. (30) as the basis for our analysis, as this parcellation is widely used and has shown superior functional homogeneity compared to other leading approaches (30). Our study focused on the 100-region parcellation, organized into 7 networks (s100) but we repeated the analyses using the 200-region variant to check the robustness of the results (see supplementary material). Regions were screened for low BOLD signal intensity, with a method adapted from Brown et. al. (61). Specifically, we found the elbow of the BOLD signal distribution, given by the largest decrease in pair-wise differences of the mean BOLD signal of each region. This was used as a cut-off for signal dropout and regions with lower signal than the cut-off were considered to have signal dropout. Regions that were found to have signal dropout in over 5% of subjects were excluded before analysis. For the s100 atlas, 15 regions were excluded; for the s200 atlas, 16 regions were excluded from further analysis.

To derive individually-tailored parcellations, we used the Group Prior Individualized Parcellation (GPIP) model (27), which relies on a Bayesian formulation with two priors: one based on group FC and one that drives individualized parcel boundaries. The former uses a group sparsity constraint to represent FC between parcels, which allows the model to maintain comparability between subjects. The latter uses a Markov Random Field in the form of a Potts model to label the set of parcels and maximize the FC homogeneity within each parcel based on individual data. This model allows for comparability between subjects, as it maintains the same areas and labels for every individual while capturing the variability in the shape and size of each parcel to best estimate each subject’s functional regions. Individualized parcel borders were optimised across 20 iterations, starting with the group-based Schaefer atlas and iteratively alternating between updating individual borders and the group FC prior. Further details are provided by in Chong et al. (27).

For both group-based and individualized parcellations, mean timeseries were extracted for each region in the s100 and s200 atlases using each individual’s spatially normalized and denoised functional data. Product-moment correlations were then estimated for every pair of regional time series to generate FC matrices. We only consider cortical areas here as, to our knowledge, methods for developing individualized parcellations for subcortical and cerebellar regions have not yet been developed.

#### Parcellation homogeneity and variability

We compared the within parcel functional homogeneity of the group-based and individualized parcellations as per prior work (27,30). We calculated the average FC between all pairs of vertices in a given parcel *i*, denoted *FC*_*i*_. Then, parcellation homogeneity *H* was normalised by parcel size as follows:

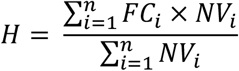

where *n* is the total number of parcels in the parcellation and *NV* is the number of vertices in the *i*^*th*^ parcel. This analysis was done out of sample, on functional scans from the second run (PE=PA) with parcellations generated for scans from the first run (PE=AP).

#### Case-control differences in inter-regional functional coupling

We assessed how parcellation type influences FC differences between patients with psychosis and healthy controls in three ways. First, we examined the distribution of unthresholded *t*-statistics obtained at each edge using a general linear model to quantify group-mean differences between patients and controls while controlling for age, sex, and test site. The contrast was specified such that a larger t-statistic indicated lower FC in patients, compared to controls. The effect of parcellation type was evaluated using a shift function test on these distributions (35) to evaluate whether differences between parcellations were restricted to specific quantiles of the *t*-statistic distributions (rather than just comparing the means of these distributions). The shift function computes the difference in value of the 9 deciles of the distributions. For inference, it computes the 95% CI associated with each decile difference, based on a bootstrap estimation of the standard error of each decile, controlling for multiple comparisons, via the Hochberg’s methods. This analysis thus allowed us to determine whether parcellation type preferentially affected results for edges that showed small, moderate, or large case-control differences.

Second, we compared thresholded results obtained with the Network Based Statistic (NBS) (36). NBS is an adaptation of cluster-based statistics for network data. A primary threshold of *p* = 0.05, uncorrected, was applied to the matrix of *t*-statistics obtained using the general linear model described above. The sizes of the connected components of the resulting network (in terms of number of edges) were then estimated. In this context, the connected components represent sets of nodes through which a path can be found via supra-threshold edges. The group labels (patients and controls) were permuted 5000 times and the previous steps were repeated. At each step, the size of the largest connected component was retained, resulting in an empirical distribution of maximal component sizes under the null hypothesis. The fraction of null values that exceeded the observed component sizes corresponds to a family-wise corrected *p*-value for each component. By performing inference at the level of connected components rather than individual edges, the NBS results in greater statistical power than traditional mass univariate thresholding methods (36). This analysis was repeated for each parcellation type (i.e., group-based and individualized) and scale (i.e., s100 and s200). Differences between significant component sizes observed using the two parcellation methods were then estimated and evaluated with respect to the differences between null component sizes estimated for the two approaches.

Finally, we examined how parcellation type affects case-control differences at the level of 7 canonical networks. We considered the control network; the default mode network; the dorsal attention network; the limbic network; the salience/ventral attention network; the somatomotor network; and the visual network using the seven Yeo network assignments associated with the s100 and s200 atlases (23). Specifically, we quantified the number of edges in the significant NBS component that fell within and between these seven networks. We examined both raw edge counts and counts normalized for the size of each network/network pair and quantified the correlation between the resulting network-level matrices obtained for each parcellation type.

## Data Availability

All data produced in the present study are available upon reasonable request to the authors

