## Supplementary Material for "The effect of using group-averaged or individualized brain parcellations when investigating connectome dysfunction: A case study in psychosis"

**Supplementary materials**

**Spatial and functional properties of group-based vs individualized parcellation**

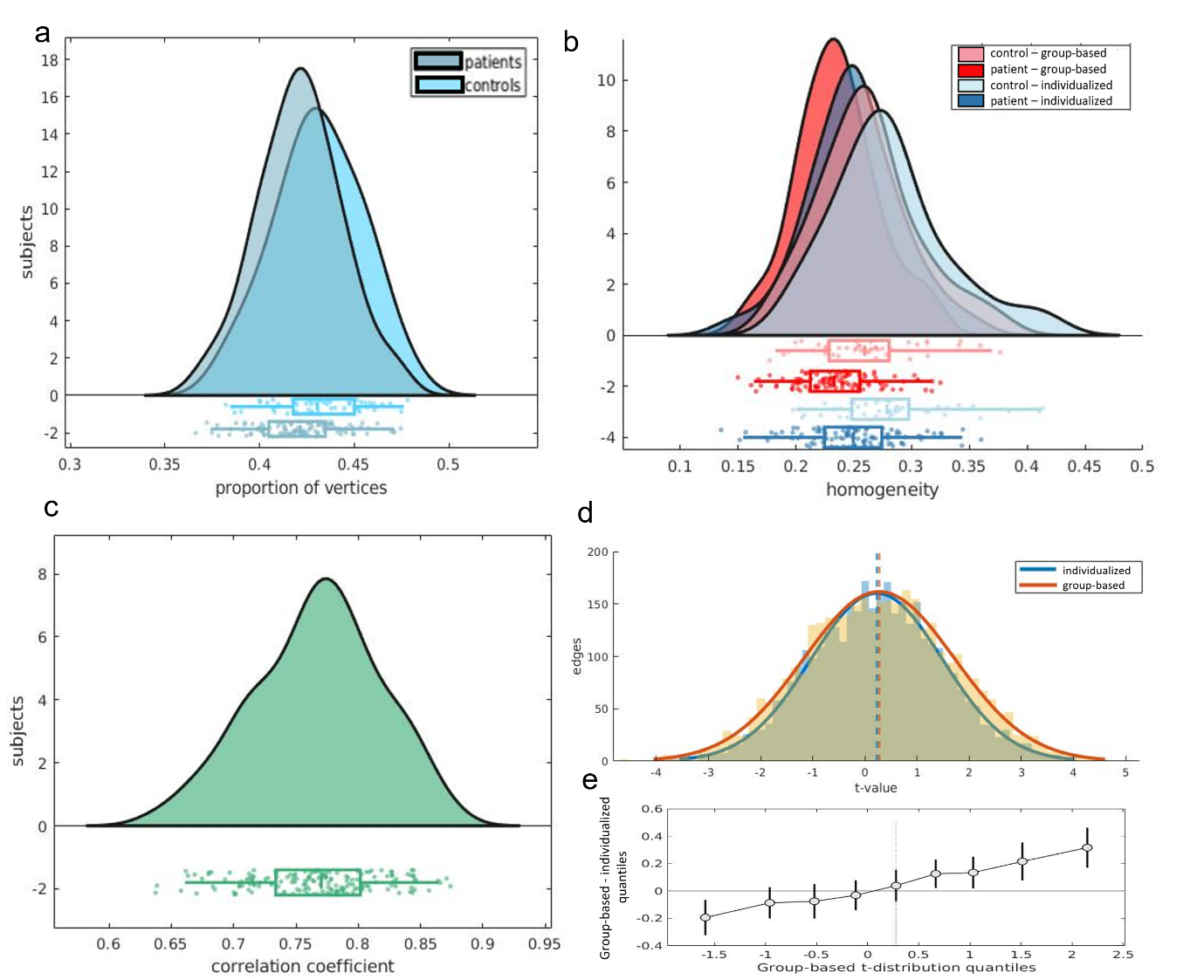

**Figure 1 - Spatial and functional properties of group-based vs individualized parcellation, based on s100 GSR. a** The proportion of vertices changed for controls $\left( M\left( SD \right)=0.431\left( 0.023 \right) \right)$and for patients $\left( M\left( SD \right)=0.421\left( 0.022 \right) \right)$. There were slightly more vertices relabelled in controls than in patients $t\left( 165 \right)=2.448, p=0.0077 95\%CI=[0.003, 0.017]$. **b** The distribution of homogeneity scores per subject. Mean homogeneity for the group parcellation in controls was $0.261 (SD=0.04)$, and $0.281 \left( SD=0.05 \right)$for the individualized parcellation. In patients, the mean homogeneity for the group parcellation was $0.235 (SD=0.04)$ and $0.250 (SD=0.04)$ for the individualized parcellation. A two-way mixed ANOVA revealed that mean homogeneity was higher for the individualized parcellation $\boldsymbol{(}F\left( 148 \right)=234.91, p<0.0001)$ and higher in controls compared to patients $(F\left( 148 \right)=15.72, p=0.0001)$, with an interaction between parcellation type and diagnosis $(F\left( 148 \right)=4.68, p=0.032)$. **c** The distribution of the Pearson’s coefficient of correlation comparing FC matrices derived from group-based and individualized parcellation. Matrices were positively correlated and ranged between 0.637 and 0.874 (median = 0.770). **d** Distributions of $t$-values quantifying FC differences between patients and controls at each edge and for individualized parcellation $\left( M\left( SD \right)=0.224\left( 1.26 \right) \right)$ and for group-based parcellation $\left( M\left( SD \right)=0.272\left( 1.44 \right) \right)$. **e** Shift function for the two t-distributions. Each circle represents the difference between each decile of both distributions, as a function of the deciles in group-based distribution and the bars represent the 95% boot-strap confidence interval associated with the difference.

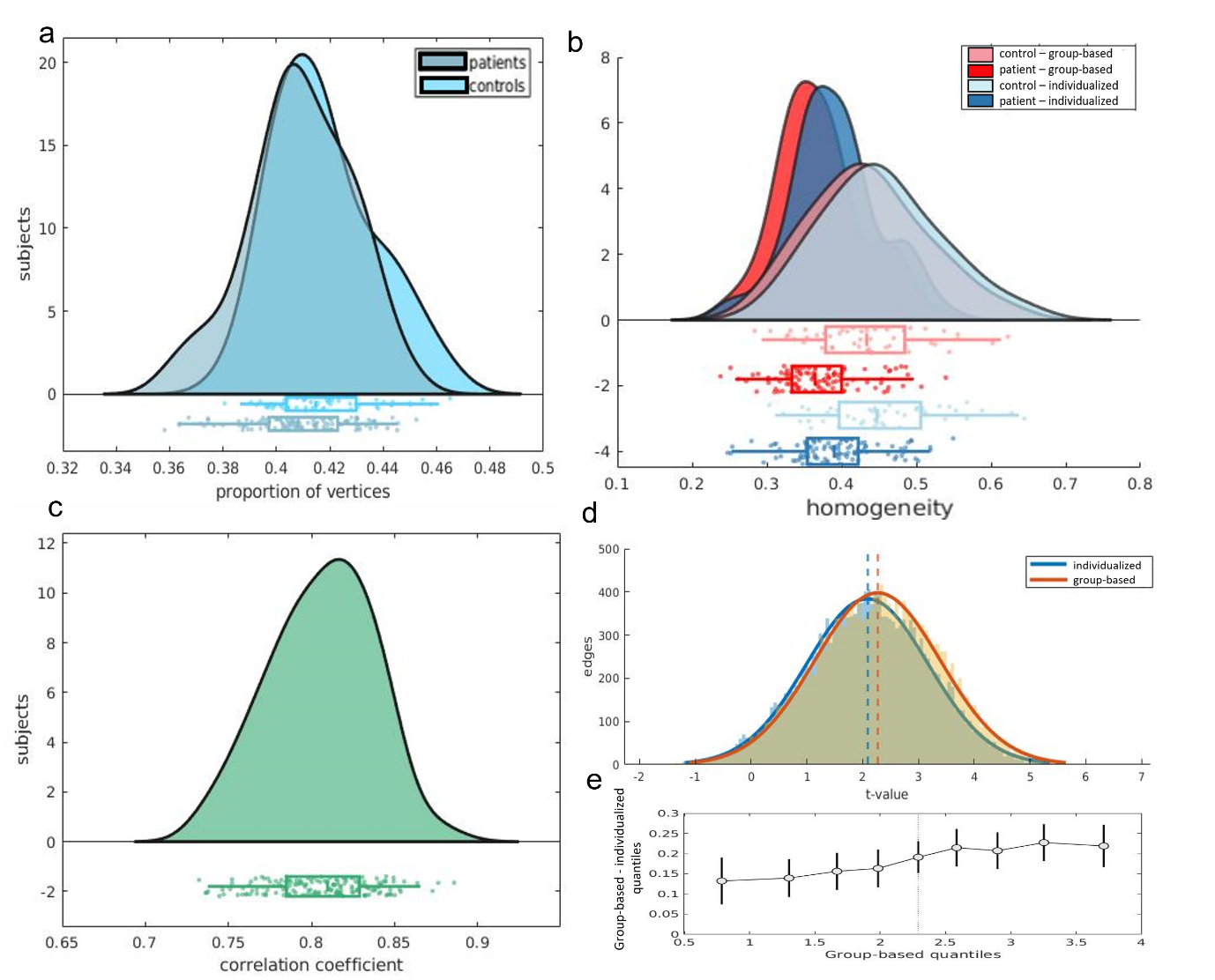

**Figure 2 - Spatial and functional properties of group-based vs individualized parcellation, based on s200. a** The proportion of vertices changed for controls $\left( M\left( SD \right)=0.418\left( 0.019 \right) \right)$and for patients $\left( M\left( SD \right)=0.409\left( 0.020 \right) \right)$. There were slightly more vertices relabelled in controls than in patients $t\left( 163 \right)=2.448, p=0.0078 95\%CI=[0.002, 0.015]$. **b** The distribution of homogeneity scores per subject. Mean homogeneity for the group parcellation in controls was $0.434 (SD=0.08)$, and $0.454 \left( SD=0.08 \right)$for the individualized parcellation. In patients, the mean homogeneity for the group parcellation was $0.371 (SD=0.06)$ and $0.392 (SD=0.06)$ for the individualized parcellation. A two-way mixed ANOVA revealed that mean homogeneity was higher for the individualized parcellation $\boldsymbol{(}F\left( 148 \right)=901.60, p<0.0001)$ and higher in controls compared to patients $(F\left( 148 \right)=29.33, p<0.0001)$, with no interaction between parcellation type and diagnosis $(F\left( 148 \right)=0.708, p=0.402)$. **c** The distribution of the Pearson’s coefficient of correlation comparing FC matrices derived from group-based and individualized parcellation. Matrices were positively correlated and ranged between 0.732 and 0.886 (median = 0.810). **d** Distributions of $t$-values quantifying FC differences between patients and controls at each edge and for individualized parcellation $\left( M\left( SD \right)=2.086\left( 1.09 \right) \right)$ and for group-based parcellation $\left( M\left( SD \right)=2.270\left( 1.13 \right) \right)$. **e** Shift function for the two t-distributions. Each circle represents the difference between each decile of both distributions, as a function of the deciles in group-based distribution and the bars represent the 95% boot-strap confidence interval associated with the difference.

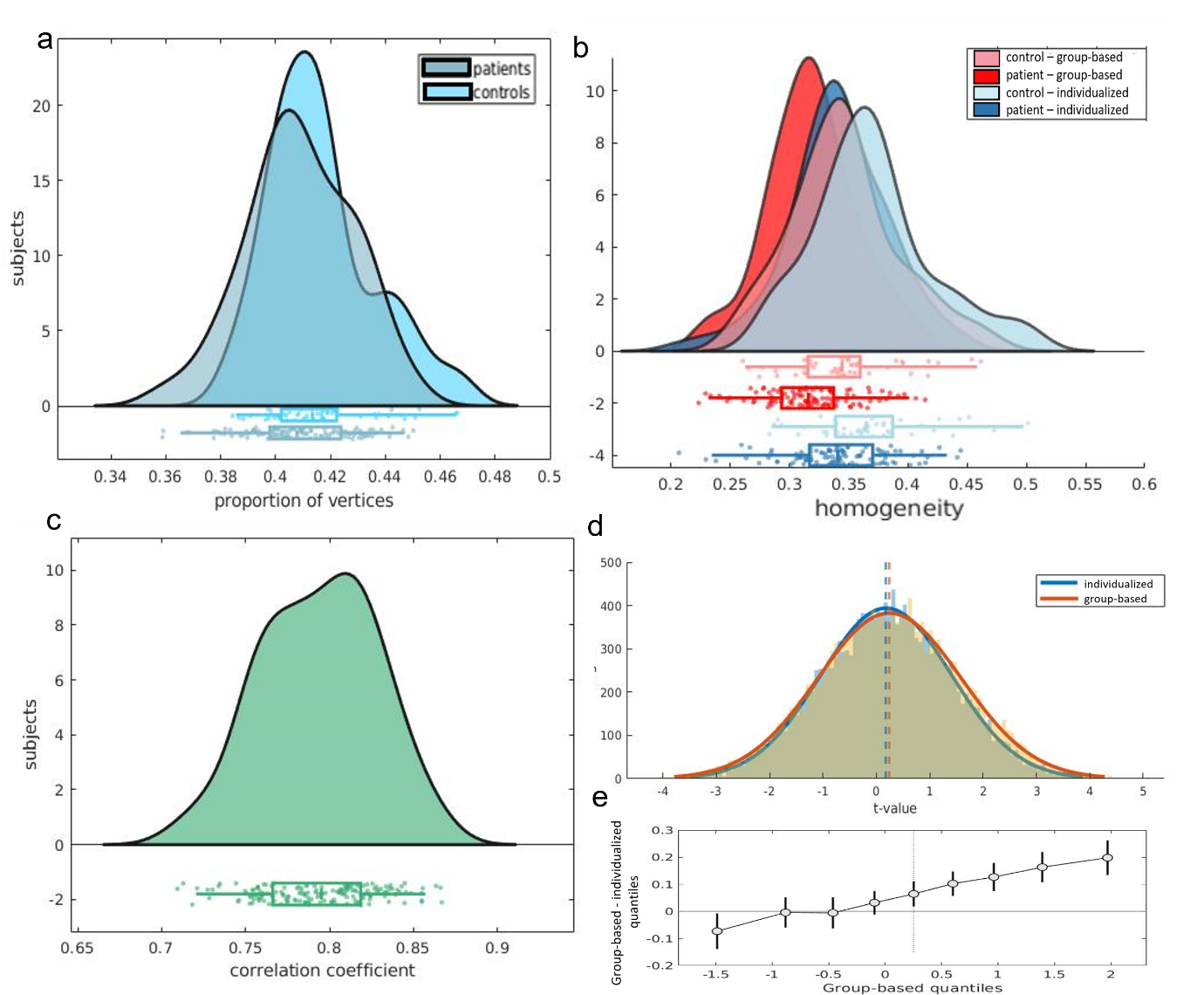

**Figure 3 - Spatial and functional properties of group-based vs individualized parcellation, based on s200 GSR. a** The proportion of vertices changed for controls $\left( M\left( SD \right)=0.416\left( 0.019 \right) \right)$and for patients $\left( M\left( SD \right)=0.409\left( 0.019 \right) \right).$ There were slightly more vertices relabelled in controls than in patients $t\left( 163 \right)=2.479, p=0.0071 95\%CI=[0.001, 0.014]$. **b** The distribution of homogeneity scores per subject. Mean homogeneity for the group parcellation in controls was $0.346 (SD=0.05)$, and $0.370 \left( SD=0.05 \right)$for the individualized parcellation. In patients, the mean homogeneity for the group parcellation was $0.318 (SD=0.04)$ and $0.341 (SD=0.04)$ for the individualized parcellation. A two-way mixed ANOVA revealed that mean homogeneity was higher for the individualized parcellation $\boldsymbol{(}F\left( 148 \right)=1040.06, p<0.0001)$ and higher in controls compared to patients $(F\left( 148 \right)=14.35, p=0.0002)$, with no interaction between parcellation type and diagnosis $(F\left( 148 \right)=0.246, p=0.621)$. **c** The distribution of the Pearson’s coefficient of correlation comparing FC matrices derived from group-based and individualized parcellation. Matrices were positively correlated and ranged between 0.710 and 0.867 (Median = 0.795). **d** Distributions of $t$-values quantifying FC differences between patients and controls at each edge and for individualized parcellation $\left( M\left( SD \right)=0.183\left( 1.23 \right) \right)$ and for group-based parcellation $\left( M\left( SD \right)=0.246\left( 1.34 \right) \right)$. **e** Shift function for the two t-distributions. Each circle represents the difference between each decile of both distributions, as a function of the deciles in group-based distribution and the bars represent the 95% boot-strap confidence interval associated with the difference.

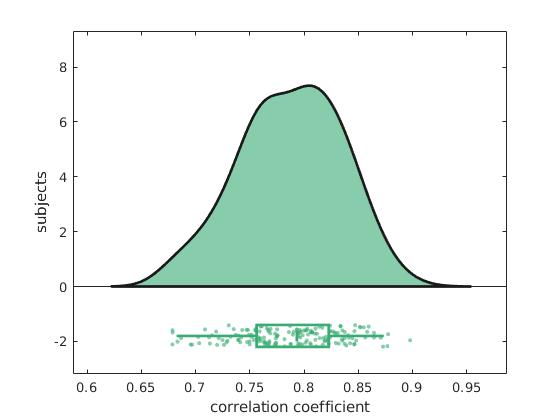

**Figure 4 – Correlation between FC matrices derived from different parcellation approaches, based on s100.** The graph shows the distribution and boxplot of the Pearson’s coefficient of correlation comparing FC matrices derived from group-based and individualized parcellation. Matrices were positively correlated and ranged between 0.679 and 0.898 (median = 0.794).

**Thresholded group differences in FC according to parcellation method**

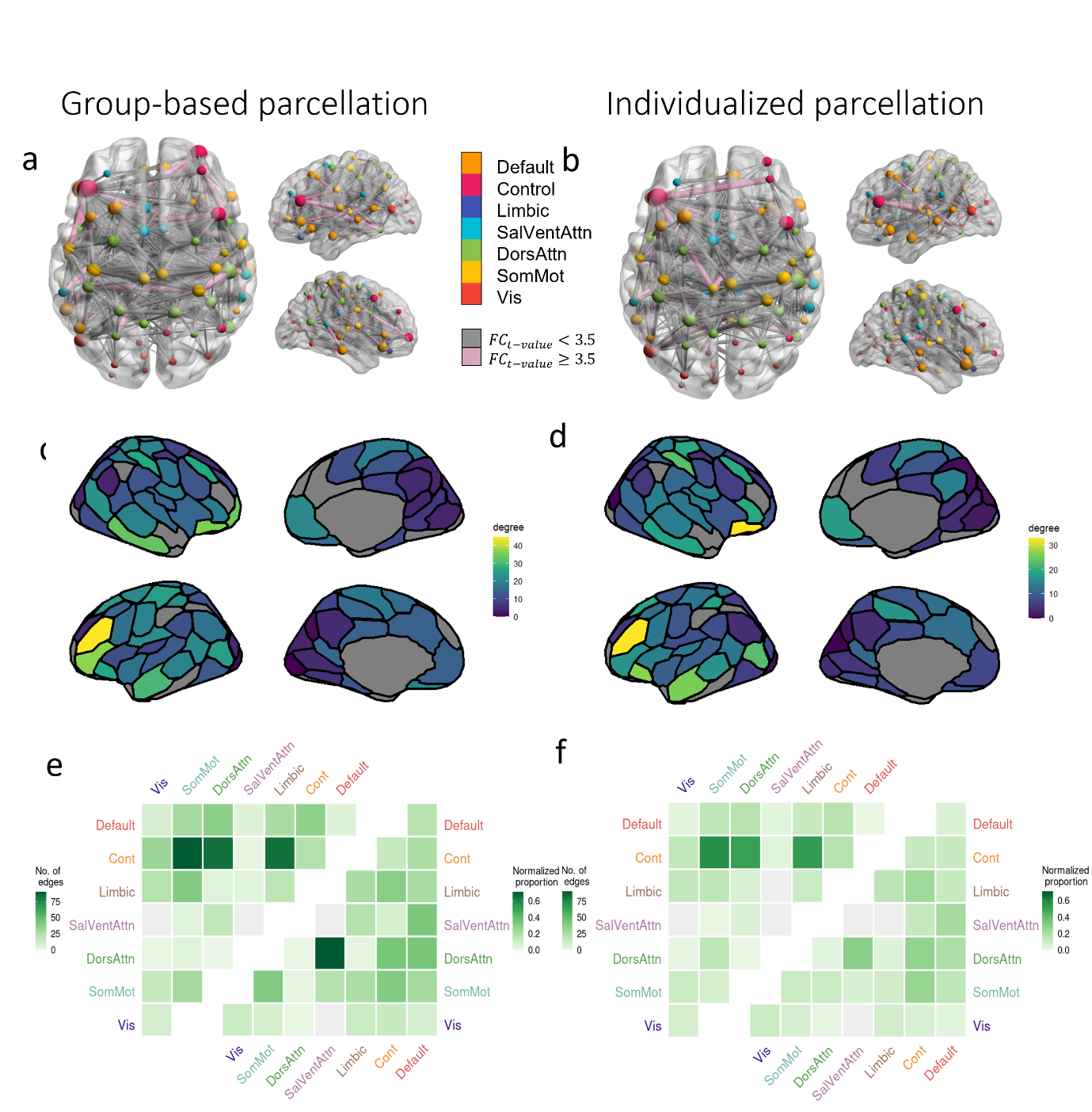

**Figure 5 – Edge-level regional and network-level case-control FC differences according to parcellation type, based on s100 GSR.** The NBS identified a single connected component as showing significant FC differences between groups using both the (**a**) group-based $(p=0.0004)$ and (**b**) individualized parcellations $(p=0.001)$. The group-based component (**a** and **c**) comprises 637 edges and the individualized component (**b** and **d**) comprises 458 edges. Panels **a** and **b** show the specific edges comprising the NBS components obtained with the group-based and individualized parcellations, respectively, with nodes colored according to network affiliation and sized by degree. Edges are sized by strength of dysconnectivity. Edges associated with a t-value < 3.5 are represented by grey lines and those associated with a t-value $\geq$ 3.5 are represented in pink. The images were created using the software BrainNet Viewer (1). Panels **a**, **c** and **e** are based on group parcellation. Panels **c** and **d** show the degree of each region in the NBS component for the group and individualized parcellations, respectively. Edges are represented by grey lines. The upper triangle of each matrix in panels **e** and **f** shows the total number of NBS component edges (raw counts) falling within and between seven canonical networks. The lower triangles show the same data normalized for network size (normalized counts). Vis – visual network; SomMot – somatomotor network; DorsAttn – dorsal attention network; SalVentAttn – salience/ventral attention network; Cont – control network; Default – Default Mode Network.

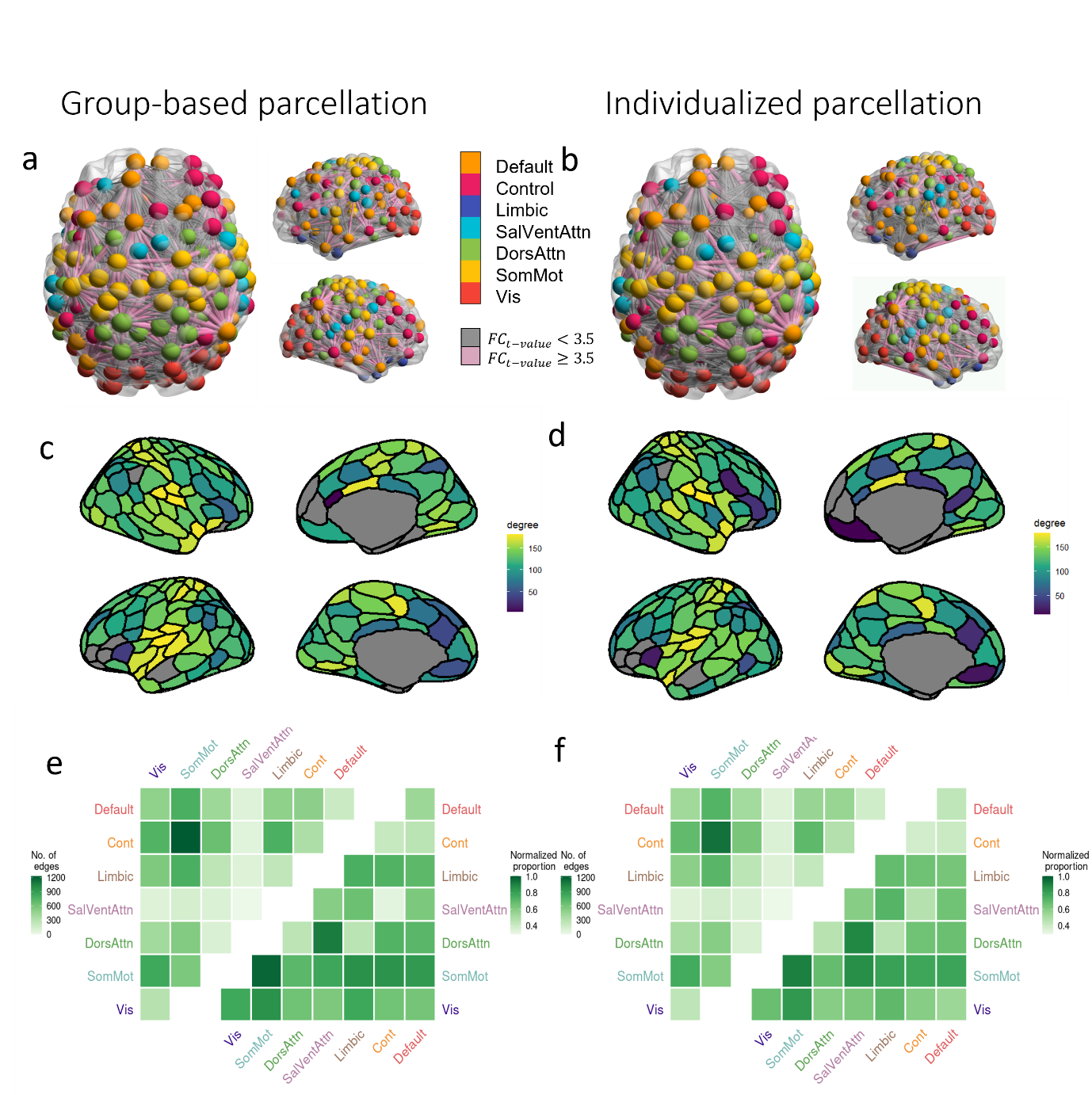

**Figure 6 – Edge-level regional and network-level case-control FC differences according to parcellation type, based on s200.** The NBS identified a single connected component as showing significant FC differences between groups using both the (**a**) group-based $(p=0.0004)$ and (**b**) individualized parcellations $(p=0$; i.e., no null value exceeded the observed estimate). The group-based component (**a** and **c**) comprises 11,852 edges and the individualized component (**b** and **d**) comprises 11,026 edges. Panels **a** and **b** show the specific edges comprising the NBS components obtained with the group-based and individualized parcellations, respectively, with nodes colored according to network affiliation and sized by degree. Edges are sized by strength of dysconnectivity. Edges associated with a t-value < 3.5 are represented by grey lines and those associated with a t-value $\geq$ 3.5 are represented in pink. The images were created using the software BrainNet Viewer (1). Panels **a**, **c** and **e** are based on group parcellation. Panels **c** and **d** show the degree of each region in the NBS component for the group and individualized parcellations, respectively. Edges are represented by grey lines. The upper triangle of each matrix in panels **e** and **f** shows the total number of NBS component edges (raw counts) falling within and between seven canonical networks. The lower triangles show the same data normalized for network size (normalized counts). Vis – visual network; SomMot – somatomotor network; DorsAttn – dorsal attention network; SalVentAttn – salience/ventral attention network; Cont – control network; Default – Default Mode Network.

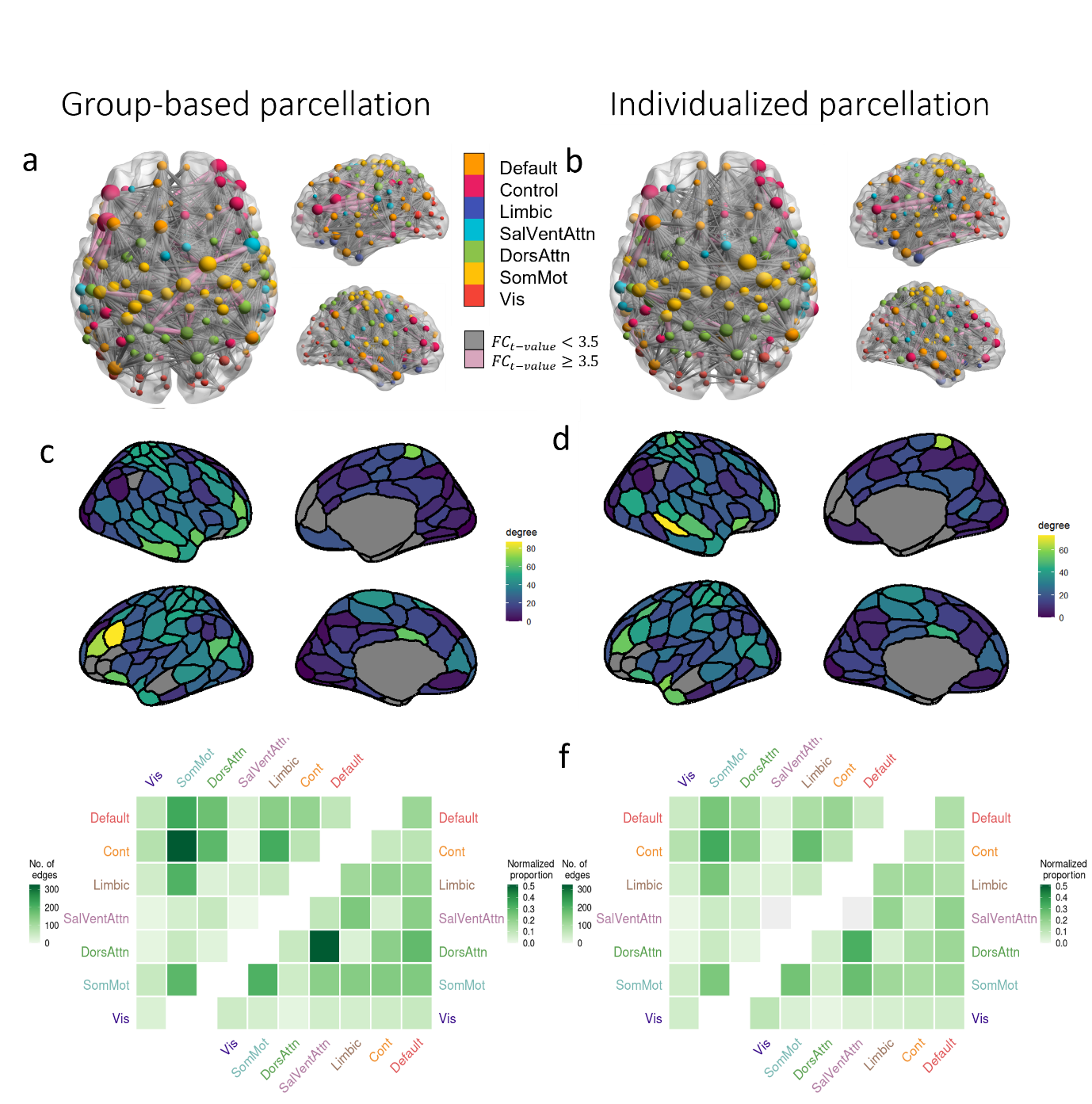

**Figure 7 – Edge-level regional and network-level case-control FC differences according to parcellation type, based on s200 GSR.** The NBS identified a single connected component as showing significant FC differences between groups using both the (**a**) group-based $(p=0.001)$ and (**b**) individualized parcellations $\left( p=0.0018 \right).$The group-based component (**a** and **c**) comprises 2,513 edges and the individualized component (**b** and **d**) comprises 2,002 edges. Panels **a** and **b** show the specific edges comprising the NBS components obtained with the group-based and individualized parcellations, respectively, with nodes colored according to network affiliation and sized by degree. Edges are sized by strength of dysconnectivity. Edges associated with a t-value < 3.5 are represented by grey lines and those associated with a t-value $\geq$ 3.5 are represented in pink. The images were created using the software BrainNet Viewer (1). Panels **a**, **c** and **e** are based on group parcellation. Panels **c** and **d** show the degree of each region in the NBS component for the group and individualized parcellations, respectively. Edges are represented by grey lines. The upper triangle of each matrix in panels **e** and **f** shows the total number of NBS component edges (raw counts) falling within and between seven canonical networks. The lower triangles show the same data normalized for network size (normalized counts). Vis – visual network; SomMot – somatomotor network; DorsAttn – dorsal attention network; SalVentAttn – salience/ventral attention network; Cont – control network; Default – Default Mode Network.

**The effects of variation in parcel size**

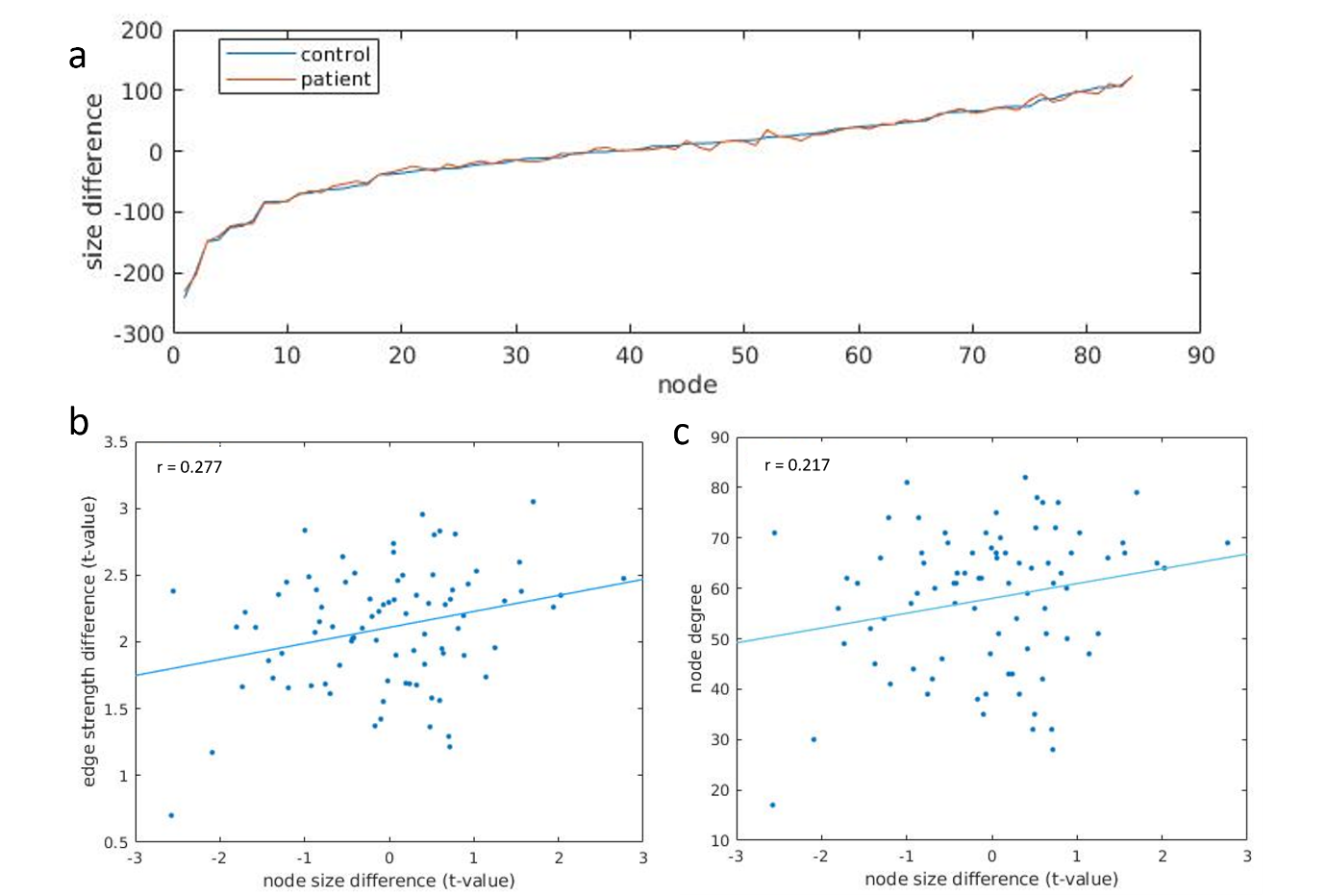

**Figure 8 – Changes in parcels size and its correlation to node degree and edge dysconnectivity, based on s100. a** The average difference in size of every region between individualized and group-based parcellation for patients and controls. Size is measured in terms of vertices and the change reported is the average size difference for controls and patients. There was no difference in parcel size changes between $( p=0.889.)$ Panel **b** shows the node size difference and average edge dysconnectivity for every region (blue dots) and the correlation between both (blue line) for individualized parcellation. Panel **c** shows the node size difference and node degree for every node (blue dots) and the correlation between both (blue line) for individualized parcellation. Correlation is given by the Pearson’s coefficient.

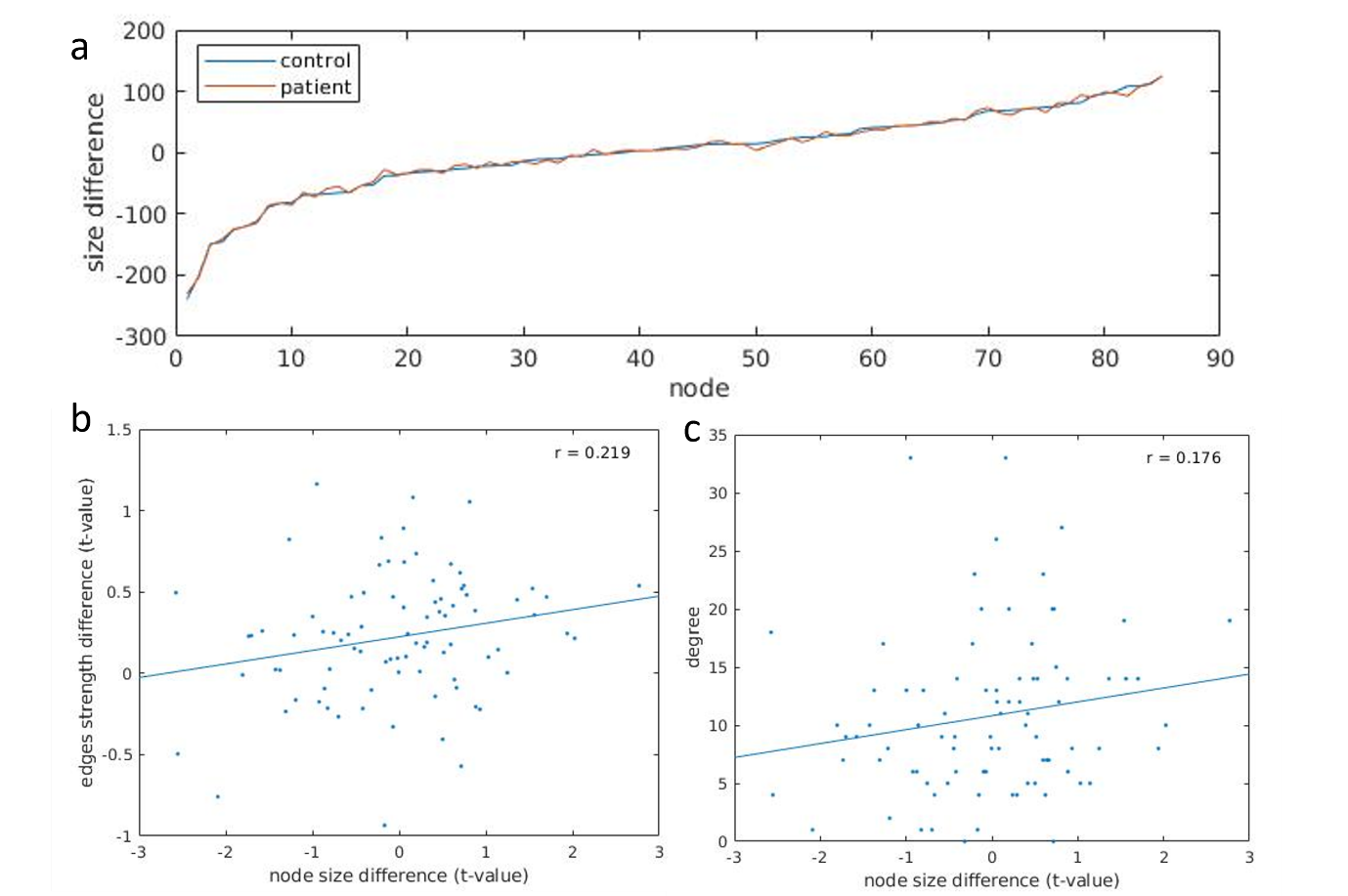

**Figure 9 – Changes in parcels size and its correlation to node degree and edge dysconnectivity, based on s100 GSR. a** The average difference in size of every region between individualized and group-based parcellation for patients and controls. Size is measured in terms of vertices and the change reported is the average size difference for controls and patients. There was no difference in parcel size changes between groups $(p=0.992).$ Panel **b** shows the node size difference and average edge dysconnectivity for every region (blue dots) and the correlation between both (blue line) for individualized parcellation. Panel **c** shows the node size difference and node degree for every node (blue dots) and the correlation between both (blue line) for individualized parcellation. Correlation is given by the Pearson’s coefficient.

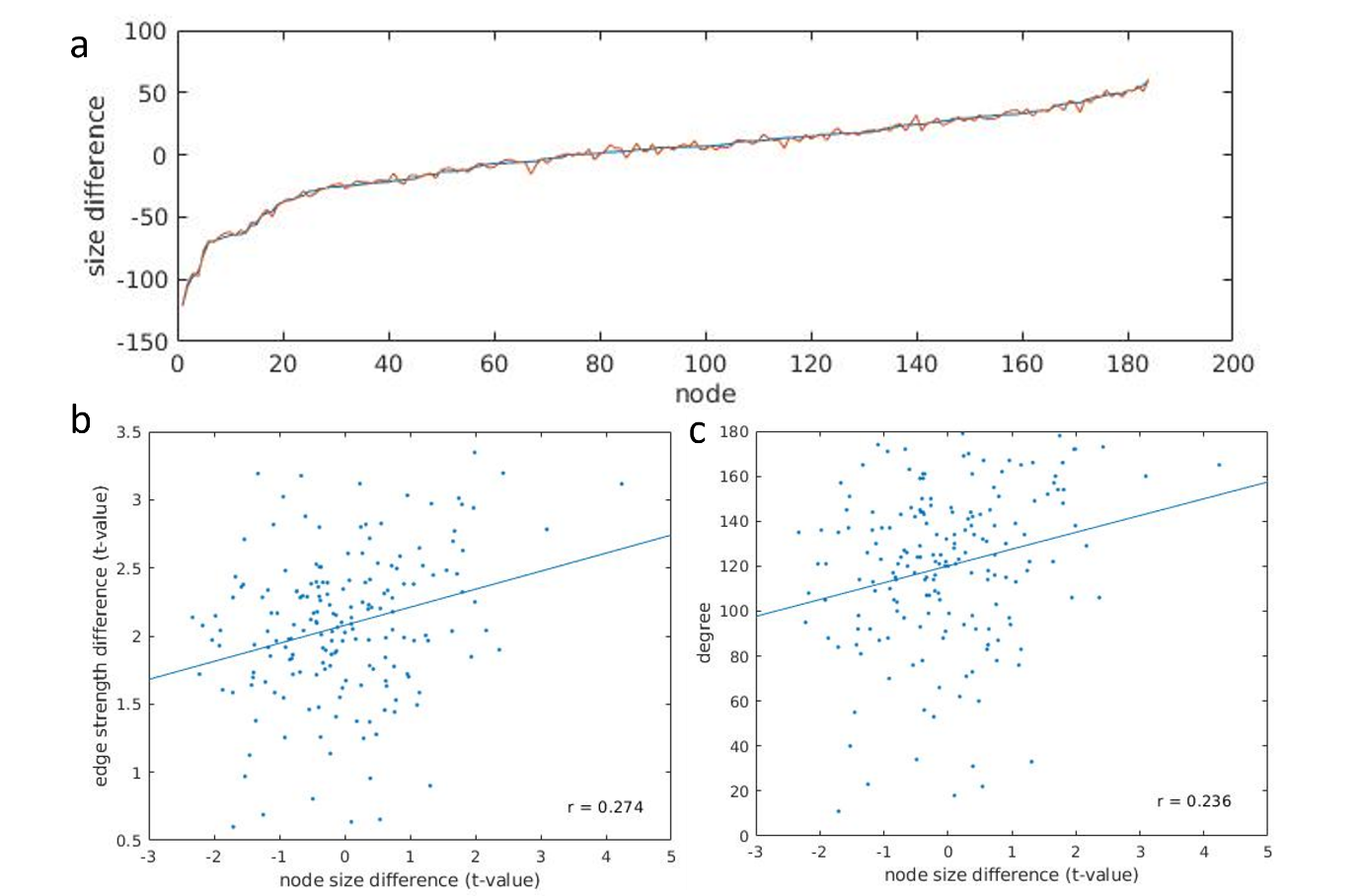

**Figure 10 - Changes in parcels size and its correlation to node degree and edge dysconnectivity, based on s200. a** The average difference in size of every region between individualized and group-based parcellation for patients and controls. Size is measured in terms of vertices and the change reported is the average size difference for controls and patients. There was no difference in parcel size changes between groups $(p=0.992).$ Panel **b** shows the node size difference and average edge dysconnectivity for every region (blue dots) and the correlation between both (blue line) for individualized parcellation. Panel **c** shows the node size difference and node degree for every node (blue dots) and the correlation between both (blue line) for individualized parcellation. Correlation is given by the Pearson’s coefficient.

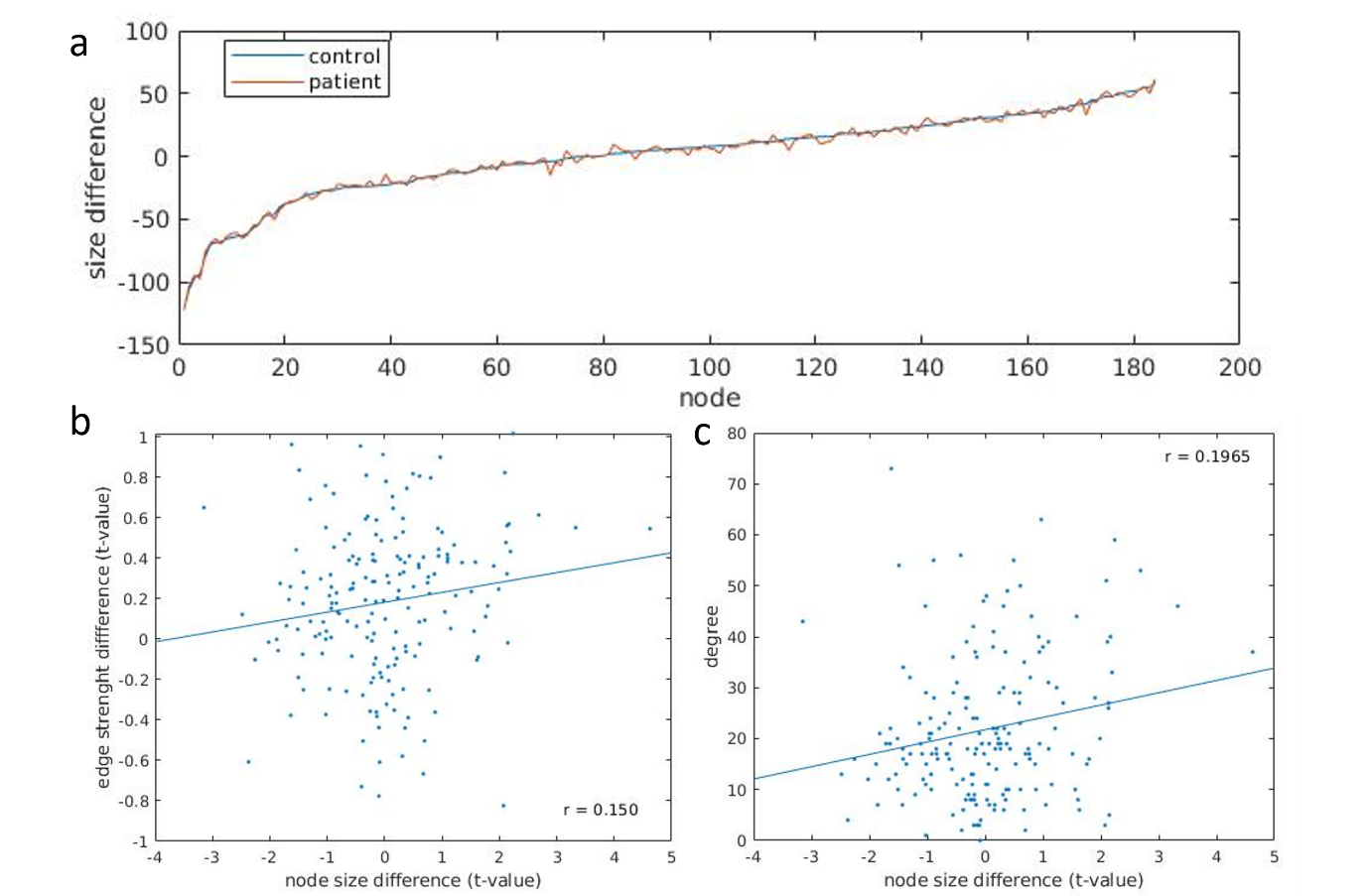

**Figure 11 – Changes in parcels size and its correlation to node degree and edge dysconnectivity, based on s200 GSR. a** The average difference in size of every region between individualized and group-based parcellation for patients and controls. Size is measured in terms of vertices and the change reported is the average size difference for controls and patients. There was no difference in parcel size changes between groups $(p=0.978).$ Panel **b** shows the node size difference and average edge dysconnectivity for every region (blue dots) and the correlation between both (blue line) for individualized parcellation. Panel **c** shows the node size difference and node degree for every node (blue dots) and the correlation between both (blue line) for individualized parcellation. Correlation is given by the Pearson’s coefficient.
